# A Prototype Thermoregulator for Prosthetic Socket Microenvironments in a Low-resourced Setting

**DOI:** 10.1101/2025.10.03.25337168

**Authors:** Daniel Opoku-Gyamfi, Christian Kwaku Agbewoavi, Zenith Yamoah, Harriet Amakye Ansah, Bismark Yao Akwetey, Kannan Govindan, Napoleon Abiwu

**Author notes:** **Correspondence:** Daniel Opoku-Gyamfi.

## Abstract

**Background:** Thermal discomfort significantly hinders consistent prosthesis use among amputees, particularly in low-resource settings like Ghana, where conventional sockets often lead to passive heat accumulation and associated skin issues.

**Methods:** A convection-based thermoregulator prototype was developed using widely available, open-source components. The system was integrated into a hard transtibial prosthetic socket with four strategically drilled ventilation holes. A rigorous bench-top experiment was conducted, employing a standardized heat gun to apply a consistent thermal load for 14 minutes, followed by a 26-minute cooling phase. Two identical setups were tested for direct comparison. Performance was evaluated and quantitative analysis of cooling rates using exponential decay models.

**Results:** The active thermoregulator demonstrated superior cooling, reducing the experimental socket temperature from its peak to below the 35°C comfort threshold in approximately 8 minutes, achieving a total reduction of 13.9°C at an average rate of 0.53°C/min over the 26-minute cooling phase. In contrast, the control group exhibited a slower, natural cooling, with a total reduction of 6.5°C at an average rate of 0.25°C/min. Quantitative analysis of cooling constants confirmed the active system’s rate (k=0.01/min) was 2.5 times greater than natural convection (k=0.004/min). These findings establish the fundamental feasibility of an active, convection-based thermoregulator for prosthetic sockets.

**Conclusion:** The prototype effectively demonstrates the ability to actively dissipate heat, offering a significant improvement over passive cooling.

## Introduction

Lower limb amputation (LLA) affects both the physical and mental well-being of individuals [1]. Globally, Lower Limb amputations account for 94.8% of all amputations [2]. In Ghana, 38.4% of older adults have at least one disability [3], of which amputation is included. The consequences of limb loss are particularly acute in low-resourced settings, where access to appropriate prosthetic care is limited [4]. Prosthetic abandonment is a major concern, often driven by prosthetic socket discomfort that negates the potential benefits of the device [5]. A primary, yet often overlooked, contributor to this discomfort is heat [6].

The tropical climate of Ghana, characterized by a high number of extreme heat days, exacerbates the physiological challenges at the human-prosthesis interface [7,8]. In Ghana, conventional prosthetic sockets are typically fabricated from laminated thermoset composites using materials like resin, hardener, and fiberglass/carbon fiber reinforcement which create an occlusive, non-breathable environment around the residual limb [9,10]. This leads to the accumulation of heat and sweat, with more than 61.7% of Ghanaian amputees reporting significant heat-related issues [11]. Physiologically, this sustained high temperature and moisture can lead to skin maceration, folliculitis, and bacterial or fungal infections, increasing the risk of skin breakdown and deeper tissue injury [6,12,13]. The resulting discomfort not only discourages prosthesis use, impacting mobility and quality of life, but can also lead to changes in residual limb volume, compromising the fit and stability of the socket and increasing shear forces on the skin [14].

A variety of technological solutions have been proposed globally to mitigate thermal distress in prosthetic sockets, including active cooling systems, air circulation devices, and advanced liners with phase-change materials [15–20]. While these innovations demonstrate a range of possibilities, there is a notable absence of published research on thermoregulatory solutions developed specifically for or within the African context. This represents a critical gap, as the direct applicability, accessibility, and cost-effectiveness of existing international technologies for users in Ghana have not been established. This study represents a foundational first step to address this technological disparity.

To begin bridging this technological gap, the primary objective of this research was to develop and evaluate a novel thermoregulator prototype for prosthetic sockets, created specifically to address the unique environmental and user needs of Ghana.

## Methods

### Design Methodology

The development of the thermoregulator prototype was guided by a structured engineering design process tailored for low-resource settings. The process began with a Problem Definition and Needs Assessment, identifying thermal discomfort as a critical, unaddressed barrier to consistent prosthesis use in Ghana, based on existing literature and regional context. This led to the establishment of key Design Requirements: the system had to be low-cost, constructed from widely available components from the open-source electronics ecosystem (e.g., Arduino), energy-efficient enough to be battery-powered, and simple enough for reliable operation and potential local repair.

During the Conceptual Design phase, various cooling mechanisms were considered. Passive solutions, such as heat sinks or phase-change materials, were deemed insufficient for the high ambient heat in the target environment. Active solutions, like Peltier modules, were rejected due to their higher cost and significant power consumption. A convection-based approach using a fan was selected as the most promising concept, offering a balance of effective cooling, availability, low cost, and simple implementation.

This conceptual framework then guided the Detailed Design and Development phase, which involved the specific selection of core components. The Arduino Uno microcontroller was chosen as the central control unit, a DHT11 sensor for temperature and humidity, and an axial fan for active cooling. This selection was made judiciously, aligning directly with the established design requirements for a low-resource setting. While alternative microcontrollers like the ESP32 offer integrated Wi-Fi/Bluetooth and potentially lower power consumption, the Arduino Uno was deliberately selected due to its unparalleled combination of low cost, widespread availability of components within the open-source electronics ecosystem (crucial for local sourcing and potential repair in Ghana), and simplicity of programming and integration. This choice specifically supports the objective of creating a solution that is not only functional but also accessible and maintainable by individuals with foundational electronics knowledge in resource-constrained environments. The simplicity of the Arduino Uno’s ecosystem, including readily available modules like the HC-05 Bluetooth module, also facilitated straightforward implementation of the desired monitoring and control features without introducing unnecessary complexity or reliance on specialized components. This fan was specifically selected based on the internal volume and dimensions of the transtibial prosthetic socket to ensure adequate air circulation. The chosen fan provides an airflow rate of 13.8 Cubic Feet per Minute (CFM), which was deemed sufficient to facilitate convective heat transfer within the enclosed socket microenvironment.

This phase also included the circuit design, software programming to create the control logic, and the physical assembly and integration of the system onto the prosthetic socket. The control logic was written in C++ using Arduino IDE (v. 2.3.3). The complete system, including the circuit schematic and control code, was first modeled and simulated using Proteus Design Suite (v. 8.13 SP0). Proteus simulation was utilized exclusively for virtual prototyping to validate the electronic connections and software logic in a simulated environment before physical assembly, specifically verifying component connectivity, circuit integrity, and the execution flow of the control algorithm. This step was crucial for iterative refinement and debugging of the electrical and logical aspects of the system, minimizing potential errors during hardware implementation. The Proteus simulation was not used to model or validate the thermal performance or airflow dynamics of the system. Once the simulation confirmed the system operated as intended, the physical assembly and integration of the components onto the prosthetic socket were performed. Crucial to the convection-based cooling mechanism was the design of the ventilation holes in the prosthetic socket. Four circular ventilation holes, each with a diameter of 0.4 cm, were strategically drilled into the anterior-lateral side of the prosthetic socket. This placement was chosen to optimize airflow, allowing the fan to draw cooler ambient air into the socket and expel warmer air, thereby creating a convective current within the microenvironment. The number and size of these holes were determined through iterative physical testing to balance effective air exchange with maintaining the structural integrity of the socket. The total cost of the components used to design the thermoregulatory system was $36.80, with the price quote of all components attached in Appendix I.

The final step of the methodology was the Performance Evaluation, detailed below, which was designed to test whether the developed prototype met the primary functional requirement of temperature reduction.

### Prototype Design and Function

A convection-based thermoregulator prototype was developed to actively cool the intra-socket environment. The system consists of a central control unit (Arduino Uno R3), a temperature and humidity sensor (DHT11), LCD screen, Bluetooth module (HC-05) and an axial cooling fan. While the DHT11 is a widely used and cost-effective sensor, it’s important to acknowledge its limited temperature accuracy of ±2°C. This was considered acceptable for the initial proof-of-concept prototype, which aimed to demonstrate the *feasibility* of active cooling within a prosthetic socket in a low-resource setting. These components with exception of the axial cooling fan and the LCD screen, were housed in a compact case attached to a transtibial prosthetic socket. The system operates automatically: when the internal socket temperature, measured by the sensor, exceeds a preset threshold, the microcontroller activates the fan to circulate air through ventilation holes drilled into the socket wall, thereby reducing the temperature via convection. The LCD screen also provide real-time feedback and the Bluetooth module provide optional manual control and data monitoring via a smartphone application. The system’s architecture is illustrated in the schematic diagram (Fig. 1) and its operational logic is shown in the flowchart (Fig. 2). The final assembled prototype is shown in Fig. 3. A detailed list of components and the full Arduino code are provided in Appendix II.

**Fig. 1.**
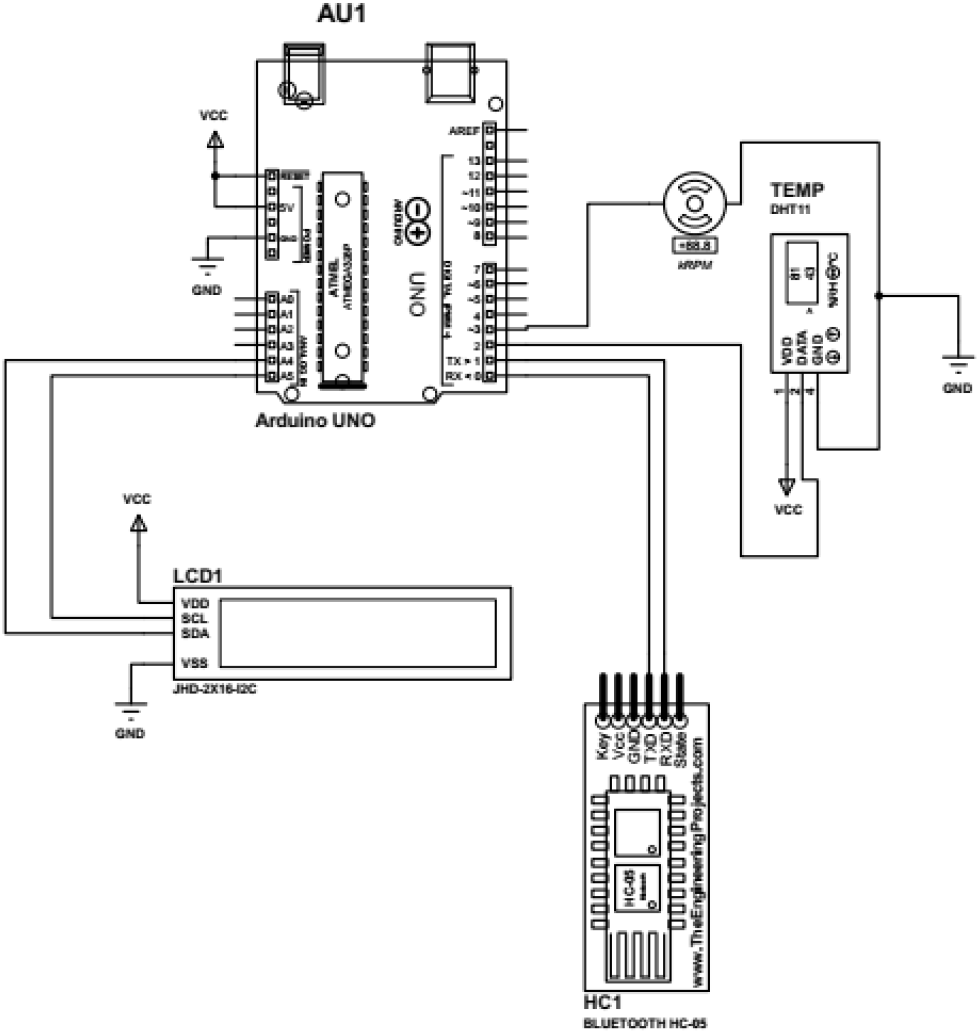
System Schematic Diagram.

**Fig. 2.**
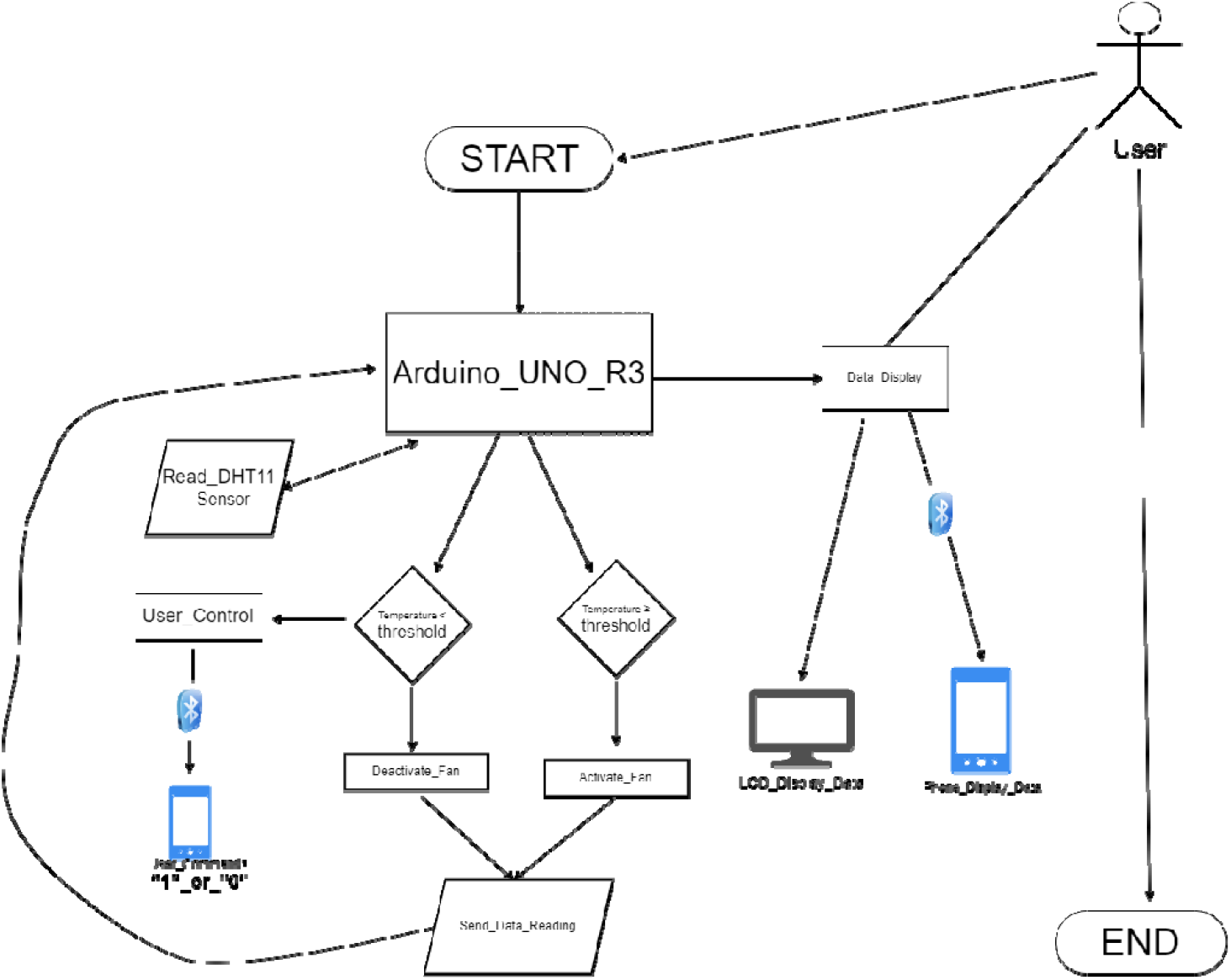
Thermoregulator Flowchart.

**Fig. 3.**
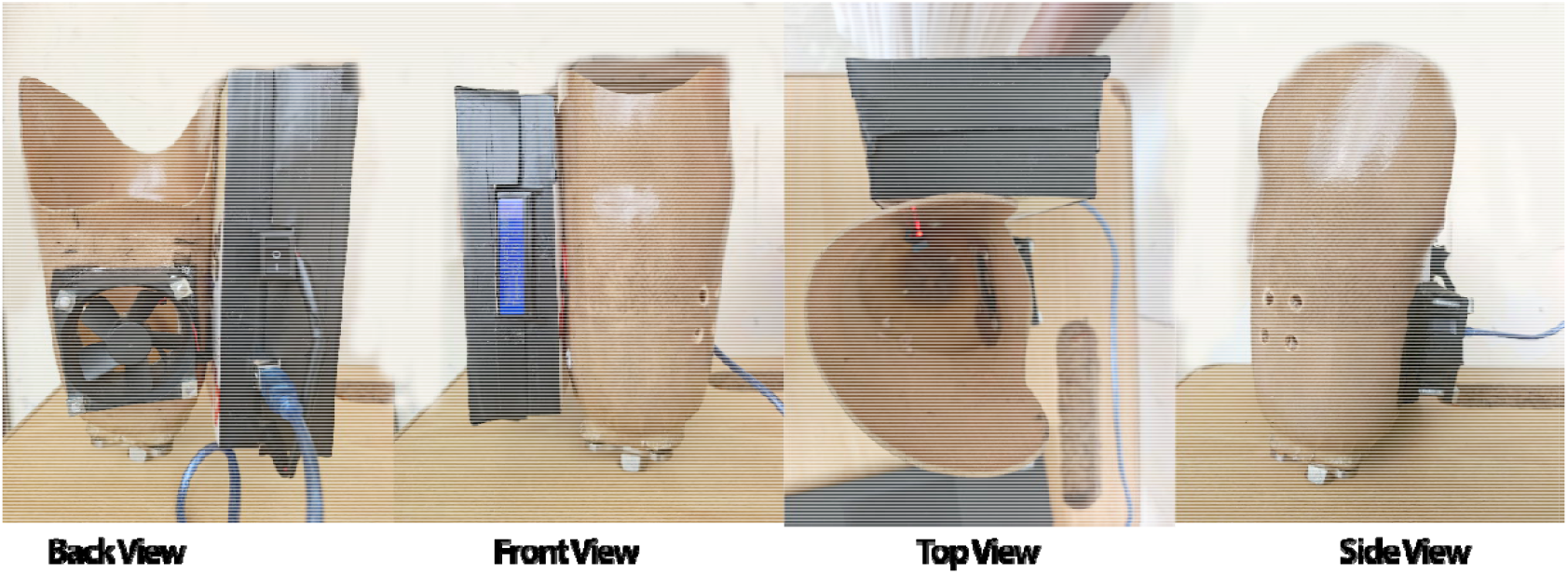
Final prototype assembly.

### Prototype Physical Characteristics

The thermoregulator system, including all electronic components and housing, has a total weight of approximately 367g, measured using a calibrated digital laboratory scale (Ohaus Scout Pro, 0.1g resolution). The scale was calibrated prior to measurement using standard 100g and 500g calibration weights to ensure accuracy. When attached to the test prosthetic socket (2.7kg), the total weight increased to 3.067kg, as verified with the same scale. The system is powered by a standard 9V alkaline battery, selected for its low cost and widespread availability.

### Performance Evaluation Protocol

A bench-top experiment was conducted to evaluate the feasibility and cooling performance of the thermoregulator prototype. The study included two distinct experimental conditions to provide a clear benchmark: an Experimental Group (with the thermoregulator active) and a Control Group (without thermoregulator).

### Experimental Setup

The assembled prototype, attached to a transtibial socket, was placed in a controlled laboratory environment. Ambient room conditions were monitored throughout the experiment, with an average temperature of 28°C (± 2°C) and relative humidity of 75% (± 5%). The DHT11 temperature sensor was affixed to the inner posterior wall of the socket to measure the internal socket environment temperature. For both the experimental and control groups, the hard socket was used without a prosthetic liner to isolate the heat transfer dynamics of the system. The DHT11 sensor used for measurement has a manufacturer-stated precision of ±2°C for temperature. Prior to testing, the sensor’s readings were verified against a standard laboratory mercury thermometer (ASTM 2C Mercury Thermometer, -5°–300°C) to ensure they fell within this specified range of accuracy. A temperature threshold of 35°C was programmed into the microcontroller as the target for thermal comfort. The threshold was chosen as a baseline for this prototype, with future work needed to optimize it for individual users.

For this initial proof-of-concept prototype, a commercial-grade, variable-temperature heat gun (Steinhel HG 2120 E) was utilized to introduce a consistent and repeatable thermal load. Prior to each experimental run, the heat gun was set to a fixed temperature setting of 130°C on the device’s dial. The heat gun’s nozzle was positioned at a fixed distance of 15 cm from the socket opening with an airflow rate of 125 L/min. A non-contact infrared thermometer (Fluke 561) was used to measure the temperature of the air at the socket’s opening, which was found to be approximately 63°C (± 5°C).

### Experimental Procedure

Two separate experimental runs were conducted, with each protocol repeated five times(n=5) to ensure reliability, and the data were averaged for analysis. The total duration of each test was 40 minutes, comprising a 14-minute heating phase and a 26-minute cooling phase.

#### Experimental Group (Forced Convection)

The socket was equipped with the complete, operational thermoregulator prototype, including the Arduino microcontroller, sensor, and fan. A heat gun was used to apply a consistent thermal load for 14 minutes until a peak temperature was reached. At the 14-minute mark, the heat source was removed, and the top of the socket was covered with a leather material to minimize passive cooling from the ambient environment. This duration allowed sufficient heat accumulation to evaluate the prototype’s cooling capacity without introducing material degradation risks. The cooling system was intentionally kept inactive during the 14-minute heating phase to establish a baseline peak temperature. The thermoregulator was then allowed to operate automatically after the 14th minute to activate the fan to circulate internal socket air.

#### Control Group (Natural Convection)

An identical socket, prepared with the same ventilation holes but with no thermoregulator or fan attached, was subjected to the exact same heating protocol. The heat gun was applied for 14 minutes, followed by its removal and the covering of the socket the same leather material. In this control run, cooling occurred solely through natural convection via the drilled ventilation holes.

### Data Collection and Analysis

For both the experimental and control groups, temperature data was logged every two minutes throughout the entire 40-minute test. Average of the logged data was used for the analysis. The primary outcome measures were the peak temperature achieved, the total temperature reduction (°C) from peak to the comfort threshold, the time to cool (minutes) and the cooling rate. To quantitatively compare the cooling performance, an exponential decay function was used to model the temperature decay curve for each group, allowing us to determine and compare the cooling constants for forced versus natural convection.

## Results

The bench-top evaluation successfully demonstrated the feasibility and core functionality of the thermoregulator prototype. The performance evaluation demonstrated the thermoregulator’s ability to significantly reduce the internal temperature of the prosthetic socket under a standardized thermal load. The results are presented from both the Experimental Group (Active Cooling) and the Control Group (Natural Cooling) to provide a direct comparison of forced versus natural convection. The successful operation of the data monitoring systems was confirmed, with real-time data correctly displayed on the Arduino IDE serial monitor and transmitted wirelessly to the mobile application (Fig. 4). The system reliably measured the internal socket temperature, automatically activated its cooling mechanism in response to thermal changes, and effectively reduced the temperature under simulated high-heat conditions.

**Fig. 4.**
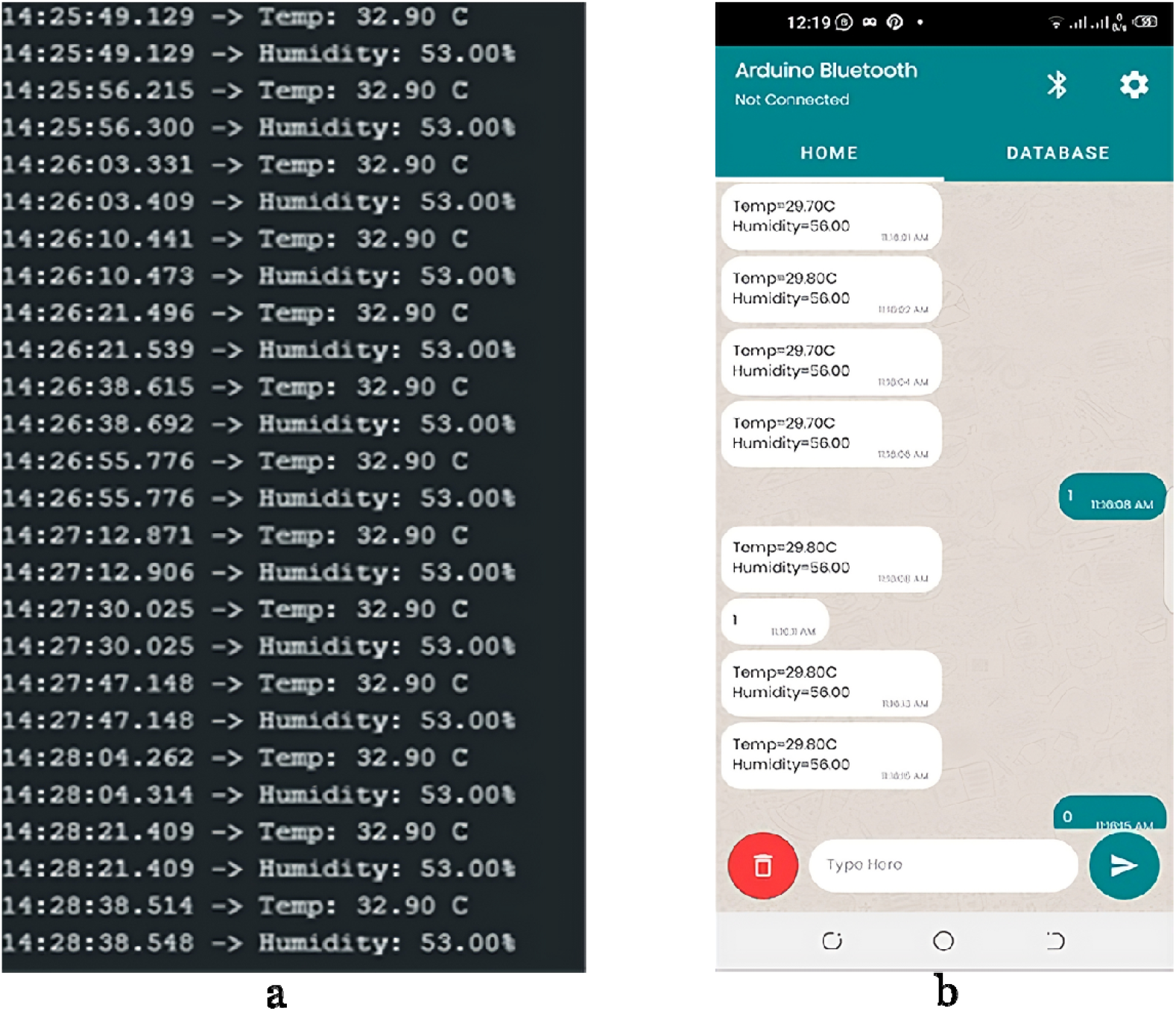
(a) Serial Monitor and (b) Mobile App Interface

### Temperature Profiles and Peak Temperature

During the heating phase (0-14 minutes), both the Experimental and Control groups showed a consistent and rapid increase in internal temperature. The average peak temperature reached at the 14-minute mark was 42.5°C for the Experimental Group and 40.3°C for the Control Group. The minor difference in peak temperature is likely attributable to slight variations in thermal mass due to the attached thermoregulator components on the Experimental Group’s socket. This difference was averagely consistent across all repeated trials. As depicted in **Fig. 5**, the temperature profiles for both the active thermoregulator and control setups demonstrated distinct behaviors during the heating and subsequent cooling periods.

**Fig. 5.**
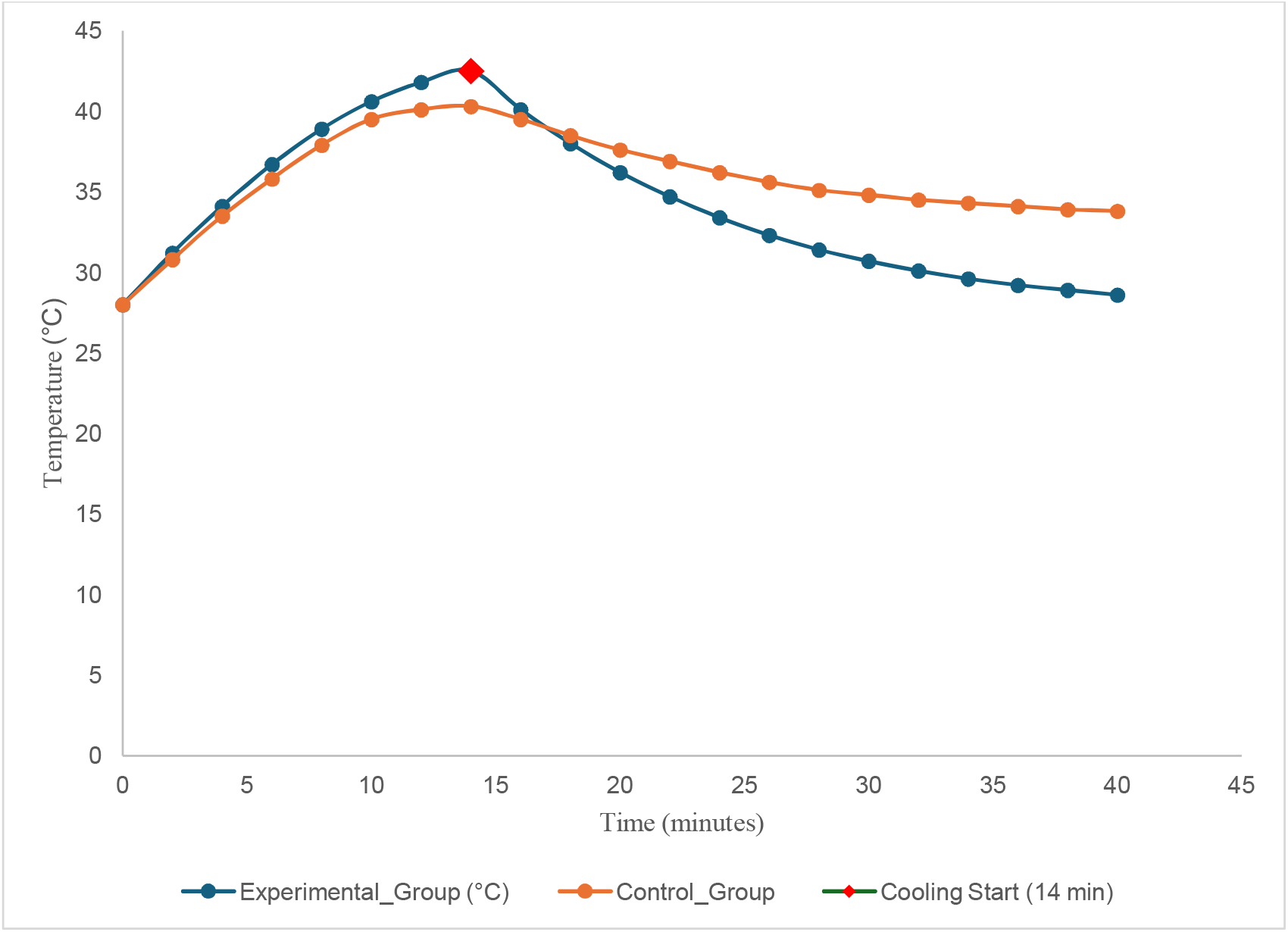
Temperature Profile of the Prosthetic Socket over the 40-Minute Experiment. This figure should show a graph with two distinct lines, one for the Experimental Group (Active Cooling) and one for the Control Group (Natural Cooling).

### Cooling Performance: Natural vs. Forced Convection

The cooling performance of the two groups was analyzed from the 14-minute mark, after the heat source was removed. **Fig. 6** provides a visual representation of the temperature reduction from the peak, with fitted exponential decay functions illustrating the difference in cooling rates. A comprehensive summary of the key performance metrics for both the experimental and control groups, including peak temperatures, total temperature reduction, and average cooling rates, is provided in **Table 1**.

**Table 1.** Comparison of Key Performance Metrics.

| Metric | Control Group (Natural Convection) | Experimental Group (Forced Convection) |
| --- | --- | --- |
| Peak Temperature (°C) | 40.3 | 42.5 |
| Time to reach 35°C (min) | ~13 (from peak) | ~8 (from peak) |
| Total Temperature Reduction (°C) over 26 min | 6.5 | 13.9 |
| Average Cooling Rate (°C/min) | ~0.25 | ~0.53 |
| Final Temperature (°C) at 40 min | 33.8 | 28.6 |

**Fig. 6.**
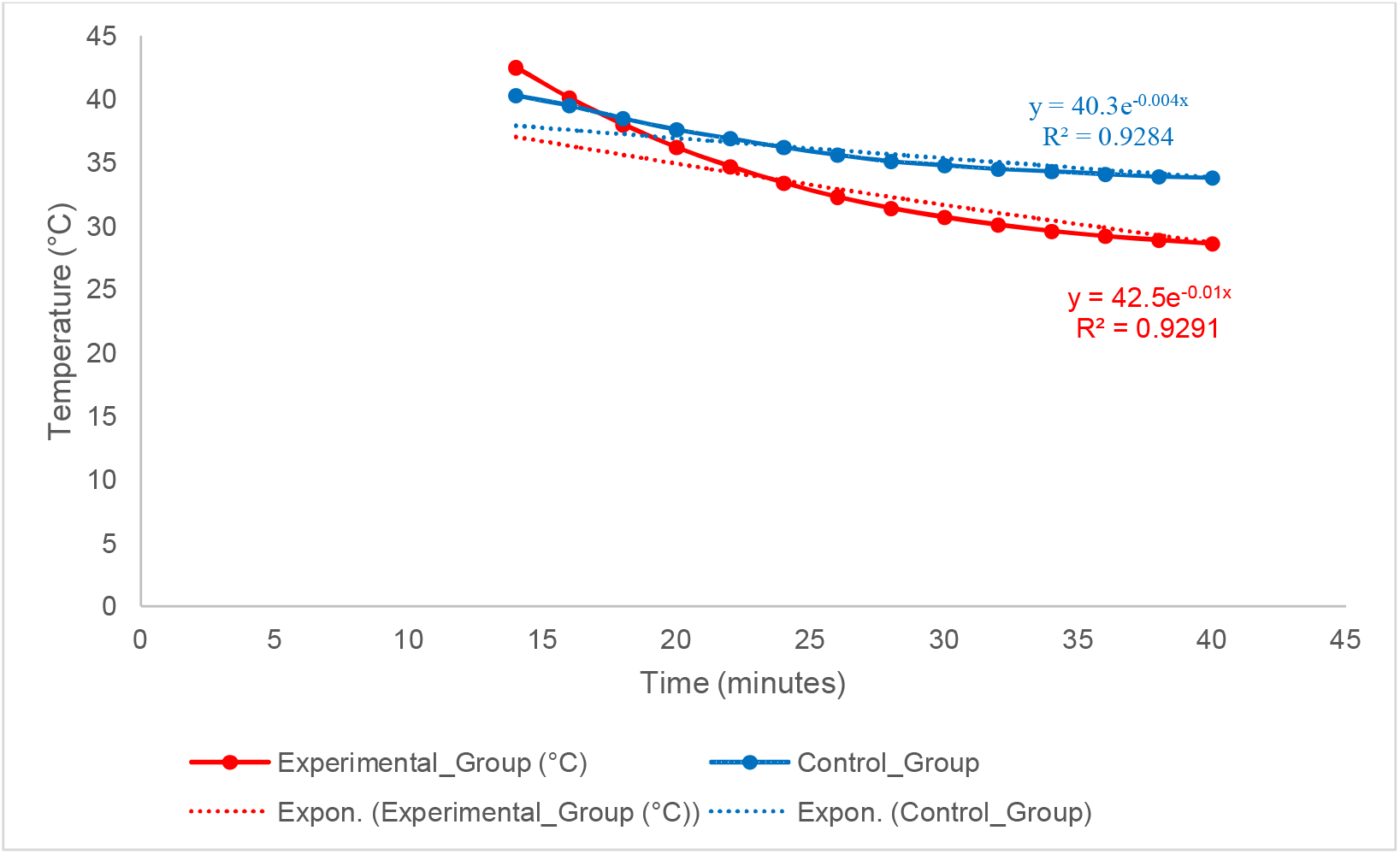
Comparison of Cooling Performance between Forced and Natural Convection. This graph illustrates the temperature decay within the prosthetic socket from the peak temperature at 14 minutes. The red curve shows the rapid cooling achieved by the active thermoregulator (forced convection), while the blue curve represents the slower, natural heat dissipation of the control group. Exponential decay functions are fitted to each dataset to model the cooling rate

In the Control Group, where cooling relied solely on natural convection through the ventilation holes, the temperature dropped from the peak of 40.3°C at 14 minutes to 33.8°C at the 40-minute mark, a total temperature reduction of **6.5°C** over 26 minutes. The system took approximately **27 minutes** (from the start of the experiment) to cool to the 35°C threshold.

In the Experimental Group, with the thermoregulator active, the cooling was significantly more rapid. The system successfully reduced the internal temperature from the peak of 42.5°C to the comfort threshold of 35°C in just **8 minutes** (from the start of cooling at 14 minutes, reaching 34.7°C at the 22-minute mark). The temperature continued to decrease, reaching a final temperature of **28.6°C** by the end of the test. The total temperature reduction from the peak was **13.9°C** over the 26-minute cooling phase.

### Quantitative Analysis of Cooling Rate

To provide a quantitative comparison of cooling performance, the temperature decay curves for both the Experimental and Control groups were modeled using an exponential decay function. This model, of the form y=A⋅e^−kx^, was fitted to the temperature data from the peak (t = 14 minutes) to the end of the test. In this equation, y represents the temperature in °C, x is the time in minutes from the start of the cooling phase (i.e., x=Time−14), A is the initial temperature at the peak, and k is the cooling constant.

The fitted equations and R-squared values for each group are as follows: **Control Group (Natural Convection):** y=40.3e^−0.004x^, R^2^=0.9284, and **Experimental Group (Forced Convection):** y=42.5e^−0.01x^, R^2^=0.9291

The cooling constant, k, is a direct measure of the rate of temperature decay. For the Control Group, the constant was k_natural_=0.004 min^−1^, while for the Experimental Group, it was k_active_=0.01 min^−1^.

A comparative analysis shows that the cooling constant of the active thermoregulator (k_active_) is 2.5 times greater than that of natural convection (k_natural_). This strong quantitative evidence, supported by high R-squared values indicating a robust fit, confirms that the forced convection mechanism provided by the fan is a highly effective method for accelerating temperature reduction within the enclosed socket environment.

## Discussion

The primary objective of this research was to develop and evaluate a low-cost thermoregulator to address heat-related discomfort in prosthetic sockets, a significant and under-addressed challenge for amputees in Ghana. The results of our bench-top evaluation robustly support our hypothesis that the developed prototype, with a total mass of approximately 367g, scan effectively reduce the temperature within a hard prosthetic socket. The 14-minute heating phase was chosen to provide a standardized, worst-case thermal stress, as this duration consistently induced temperatures exceeding physiological comfort thresholds (35°C) while remaining within safe operational limits for the sensors. This approach ensured a rigorous evaluation of the prototype’s maximum cooling efficacy. The device demonstrated a rapid and consistent cooling capability, reducing the internal socket temperature from a peak of 42.5°C to below the 35°C comfort threshold in an average of approximately 8 minutes from the start of cooling. Over the 26-minute cooling phase, the device achieved a total temperature reduction of 13.9°C at an average rate of 0.53 °C/min. More rigorously, our quantitative analysis revealed that the active thermoregulator increased the cooling rate constant (k) by 2.5 times compared to natural convection alone. These findings, while based on a bench-top model, establish the fundamental feasibility and potential clinical utility of an active, convection-based cooling approach for the local Ghanaian-African context. The ability to actively and efficiently dissipate heat, rather than merely relying on the slow process of passive heat accumulation and retention, represents a significant step forward from the status quo in conventional sockets.

A key aspect of designing for low-resource settings is affordability and accessibility. While the integration of a microcontroller and sensors may initially seem complex for such an environment, the components chosen are foundational to the global open-source electronics ecosystem [21,22]. Their mass production for hobbyist and educational markets has made them exceptionally low-cost [23]. The total bill of materials for the electronic components of this prototype is estimated to be under $40 USD. This positions the device as a significantly more accessible and financially viable pathway for local assembly and implementation when compared to specialized commercial solutions, such as phase-change material liners which can cost several hundred dollars ($539 listed on ebay), or advanced systems like the vacuum pump proposed by Bach and his colleagues with a projected retail price of $1600 [24,25]. Our design philosophy was to intentionally leverage this global economy of scale, creating a “frugal innovation” that brings modern control system capabilities to a context where they would typically be cost-prohibitive. The choice of a simplified active cooling system was a deliberate engineering trade-off between performance, complexity, and contextual appropriateness.

Our design builds upon concepts from previous research but adapts them for our specific goals. For instance, the active systems developed, demonstrated high cooling capacities (up to ∼7.0 W) using a combination of heat pipes, a heat sink, and a fan [15,18]. While powerful, these systems involved more complex and heavier components (415g), which could present challenges in cost, local sourcing, and integration into a lightweight socket [15,18]. Similarly, the “smart” system which used 16 sensors and a power-intensive Peltier thermoelectric module, offers precise control but at a level of complexity and potential cost that is less suitable for our target environment [16]. In contrast, our prototype streamlines the active cooling concept, with a total mass of 367g, by using a single fan for forced convection and intentionally omitting components like heat pipes and heat sinks. This strategic simplification minimizes complexity, manufacturing cost, and weight, aligning with our design priorities for feasibility and accessibility.

In contrast, our design offers a performance advantage over purely passive solutions. A significant temperature differential (6.4–11.1°C) was achieved through a helical cooling channel integrated within a socket wall [19], while the benefit of simple ventilation holes was demonstrated [17]. These passive methods are valuable for reducing heat buildup but are limited in their ability to actively lower temperature once it has reached an uncomfortable peak [17,19]. Our prototype incorporates the ventilation principle but enhances it with an active fan, providing more responsive and powerful cooling that can recover the thermal environment after significant heat accumulation. This hybrid approach, combining simple passive design with simple active control, appears to be a promising middle ground. Our evaluation demonstrated a total temperature reduction of 13.9°C, a value that is not only significantly larger than the 6.5°C reduction observed in our passive control group but also exceeds the reported performance range of more complex passive designs [19]. This combination of a simple passive design with a simple active control appears to be a promising middle ground in prosthetic thermoregulation. Our prototype is, to our knowledge, the first of its kind developed specifically within and for the African context, representing a foundational effort to address a locally-identified need with a globally-informed, but locally-appropriate, design. Furthermore, the customer-controlled supplementary cooling via Bluetooth and a smartphone app further enhances user preferences and flexibility, with real-time monitoring of parameters providing a significant enhancement to the user experience.

## Limitations

It is crucial to interpret the promising findings of this bench-top evaluation within the context of the study’s limitations.

A primary limitation of the current study is that the evaluation was conducted in a controlled, non-ambulatory bench-top setting. This setup, while ideal for isolating and quantifying the prototype’s cooling performance, does not account for complex variables present in clinical use, such as metabolic heat generation and perspiration from a user’s residual limb. Therefore, the cooling rates observed in this study may not be directly generalizable to a dynamic, real-world clinical application.

A second, and major, limitation is the absence of a prosthetic liner in the experimental setup. In clinical practice, the interface between the residual limb and the socket is mediated by a liner, which typically has low thermal conductivity and acts as an insulator. The fan’s forced convection primarily cools the air within the socket’s microenvironment. Therefore, the observed temperature reduction may not directly translate to the same degree of cooling at the skin-liner interface, which is the ultimate determinant of user comfort.

Furthermore, this evaluation revealed several practical challenges that must be addressed in future iterations. The addition of 367g to the prosthesis is a non-trivial weight that could affect user gait and comfort, highlighting the need for a more lightweight design. The observed battery life of approximately one hour from a standard 9V alkaline battery is also insufficient for all-day use, underscoring the necessity of a more robust power management system and a higher-capacity rechargeable power source. Finally, while the single axial cooling fan provides effective targeted cooling with an airflow of 13.8 CFM, this may lead to non-uniform temperature distribution and localized cooling, posing a potential risk of soft tissue irritation.

Based on these limitations, several key directions for future work emerge. The immediate next step must be to conduct a clinical trial with human participants to evaluate the device’s real-world effectiveness when used with common prosthetic liners (e.g., Pelite, silicone). This will require incorporating skin-temperature sensors at the liner interface to quantify the direct physiological benefit. Such a study would also provide invaluable subjective feedback on the device’s weight, noise level, and overall impact on comfort. Future design iterations should focus on optimizing the system for weight reduction and integrating a more efficient, rechargeable power source. Additionally, components such as Pulse Width Modulation (PWM) should be integrated to allow for controlled fan speed and more precise thermal regulation, mitigating the risk of non-uniform cooling.

## Conclusion

This study successfully demonstrates the feasibility and efficacy of a novel, low-cost prosthetic socket thermoregulator. Our bench-top evaluation provides a strong proof-of-concept, showing the device’s ability to reduce intra-socket temperature from a peak of 42.5°C to a comfort threshold of 35°C in just 8 minutes from the start of cooling. This rapid cooling performance, quantified by a cooling rate constant that is 2.5 times greater than that of natural convection, validates the effectiveness of our active, fan-driven approach. Our hybrid design, with a total mass of 367g, strategically combines simple passive ventilation with active forced convection to provide a context-appropriate solution for heat-related discomfort. While further development is required to address the limitations identified, such as in-vivo testing with prosthetic liners and optimizing battery life, this research establishes a critical framework for developing accessible assistive technologies in Ghana and across Africa. This work represents a pioneering effort with the potential to significantly improve the quality of life for amputees in hot climates worldwide by providing a foundational, locally-informed design with real-time monitoring and user-controlled features.

## Author Contributions

All authors reviewed and approved the final version of the manuscript. Daniel Opoku-Gyamfi: Conceptualization of the main idea, programming and coding of the Arduino microcontroller, methodology design, conducted the investigation, performed the formal analysis, and wrote the original draft of the manuscript. Christian Kwaku Agbewoavi, Zenith Yamoah, and Harriet Amakye Ansah: Contributed to the investigation, data collection, and validation of the experimental results. Bismark Yao Akwetey: Contributed to the methodology and data analysis. Kannan Govindan and Napoleon Abiwu: Provided supervision and contributed to the critical review and editing of the manuscript.

## Data Availability

The datasets used and/or analyzed during the current study are available from the corresponding author on reasonable request.

## Funding

No funds, grants, or other support was received.

## Declarations

We confirm that this work is original and has not been published elsewhere, nor is it currently under consideration for publication elsewhere.

## Conflict of interest

We have no conflicts of interest to disclose.

## Notes

### Competing Interest Statement

The authors have declared no competing interest.

### Funding Statement

This study did not receive any funding

